# Soft Temporal Scoring Using a Foundation Model: Optimal Frame Selection for Improved ONSD Measurement in Ultrasound Videos

**DOI:** 10.64898/2026.09.21.26362872

**Authors:** Mahad Ali, Luis Marin Castaneda, Chunshuang Wu, César E. Escamilla-Ocañas, Mohammad Hirzallah, Laura J. Brattain

## Abstract

Medical ultrasound is a portable, non-invasive, and cost effective imaging modality that is particularly well suited for resource-limited settings. Optic nerve sheath diameter (ONSD) measurement from ultrasound is used as a point-of-care assessment tool for intracranial pressure (ICP), which is associated with several neurological conditions. However, valid measurement depends on selecting a frame in which the optic nerve sheath is clearly visible. Manual selection requires a high level of expertise. To enable the use of ONSD in low resource settings by medical personnel of all levels, we present a sparsely supervised AI framework that scores each frame of an ONSD ultrasound video and selects the optimal frame for ONSD assessment. Per-frame embeddings from an ultrasound foundation model (USFM) are passed to a lightweight temporal head and trained using Gaussian soft labels, which assign graded targets around labeled key frames, rather than hard binary (0/1) per-frame targets. We evaluate the model performance using subject-level cross-validation on 18 subjects and 323 ultrasound videos spanning nine acquisition sweep types. Top-k frame selection is used as a metric for direct comparison against the baselines. While only requiring sparse labels, our method can identify an optimal frame in 82.2% of the evaluated sweeps, exceeding the strongest training-free baseline (49.2%) and hard-label classification (71.9%).

## 1 Introduction

Accurate measurement of the Optic Nerve Sheath Diameter (ONSD) from Ultrasound (US) videos is useful for estimating Intracranial Pressure (ICP) [9]. ICP is correlated with conditions such as Traumatic Brain Injury (TBI), strokes, hemorrhages, and central nervous system infections. However, measuring ONSD from US videos is a challenging task that requires a high level of expertise. To obtain good measurements, we need to select optimal frames from US videos where the optic nerve sheath and other landmarks are clearly visible and the optic nerve is perpendicular to the globe [4]. In this paper, we refer to these optimal frames as key frames. Key frame selection for ONSD measurement suffers from high variability because the angles and locations of the US probe on the eye can differ significantly across operators of varying skill levels. The scanning speed can also differ significantly. For example, two US videos of the same subject can look different depending on whether the sweep motion was lateral translation, vertical translation, or rotation.

Automated key frame selection can reduce the time and cost of manual selection while potentially improving consistency. This can expand access to rapid, noninvasive use in low-resource and austere settings where neuroimaging and specialized expertise are scarce. Prior approaches to frame selection in biomedical video, discussed in Section 2, have largely relied on the segmentation of anatomical structures, hand-crafted templates, or hard per-frame classification, in which each frame is assigned a binary target (1 for a labeled key frame and 0 otherwise). Each is demanding in a low-resource setting: segmentation requires dense pixel-level labeling that is costly to obtain and difficult for the small, low-contrast structures of the eye; hand-crafted templates are sensitive to anatomical heterogeneity; and hard labels impose abrupt binary boundaries between visually similar adjacent frames and provide no graded signal for ranking frames within the valid region. This can encourage overconfident predictions and overfitting to ambiguous label boundaries.

We therefore formulate key frame selection as sparsely supervised temporal frame-quality scoring using Gaussian soft labels, which assign continuous targets that peak at labeled key frames and decrease smoothly for nearby frames. We hypothesize that this graded supervision better reflects the similarity between neighboring ultrasound frames, while a lightweight LSTM temporal head over frozen ultrasound-foundation-model features can limit the number of trainable parameters to a reasonable level. Our contributions are as follows:

- We formulate ONSD key frame selection as sparsely supervised temporal frame-quality scoring and adapt Gaussian soft labels to provide graded supervision around expert-labeled frames.
- We instantiate this with a lightweight temporal head over frozen ultrasound foundation model (FM) features. Using FM can drastically reduce the amount of ONSD data needed.
- We evaluate on 18 subjects and 323 ultrasound video scans spanning nine controlled acquisition sweep types. Clinically-aligned metrics include top-k selection, which is used for locating a valid/optimal frame, and the distance from the selected frame to the nearest key frame. The method selects a valid frame in 82.2% of evaluated sweeps, compared with 71.9% for the hard-label model and 49.2% for the strongest training-free baseline.

## 2 Previous Work

### Frame selection in medical ultrasound

Prior methods often derive frame quality from anatomical segmentation. Wen et al. [12] combine segmentation with rule-based filtering to identify ONSD key frames within an ICP-grading pipeline. Ciusdel et al. [1] use segmentation followed by spatiotemporal classification to identify end-diastolic frames, while Li et al. [8] score ONSD frames using localized anatomy and a hand-designed echogenicity template. These approaches require dense pixel-level supervision and, in the template-based setting, manually designed anatomical priors. Lee et al. [7] instead compare segmented liver anatomy with an expert-selected anchor frame using image-similarity metrics. This avoids direct key frame classification but requires a reference frame at inference and assumes that a single anchor represents the range of valid views. A separate family treats frame selection as hard per-frame classification. In contrast, our method predicts graded frame quality directly from sparse frame labels.

### Soft labels for localization

Payer et al. [10] regress spatial Gaussian heatmaps for anatomical landmark localization. Wang et al. [11] extend this idea temporally by placing Gaussian targets around cardiac phase frames. We adapt this form of temporal soft supervision to ONSD key frame selection, where adjacent frames are often visually similar. Our formulation is paired with temporal scoring over frozen ultrasound foundation model features to rank frames within each sweep.

## 3 Methodology

### 3.1 Dataset and Preprocessing

Upon Institutional Review Board approval, we obtained a dataset comprising 18 subjects, both eyes, with nine acquisition sweeps per eye: an optimal ONSD view and eight controlled ultrasound video scans, each starting from the optimal view (upward/downward/leftward/rightward translation, clockwise/counter-clockwise rotation of up to 30 degrees, and upward/downward fanning). This yields 323 sweeps and approximately 45,500 frames (on average 141.0 frames/sweep). The number of labeled key frames per sweep ranges from 0 to 132 (mean 34.2), and 80/323 sweeps (24.8%) do not contain any key frames. Frames were cropped to the relevant ultrasound region, excluding probe- and device-related text and other content. We use subject-level splits; no subject appears in both the training and held-out evaluation folds. Sweeps without key frames were retained during training with all-zero targets but were excluded from localization evaluation.

### 3.2 Frame Quality Scoring Framework

We formulate key frame selection as frame-quality scoring, assigning each frame a score in [0, 1] that reflects its suitability for ONSD measurement. **Frozen feature extraction:** Each frame is independently encoded by the frozen Ultrasound Foundation Model (USFM), a Vision Transformer pre-trained on ultrasound images [6, 2], producing a *D* = 768-dimensional feature vector. USFM reduces the amount of ONSD data needed. Freezing the encoder limits task-specific learning to a small number of parameters. **Temporal head:** For a sweep of *T* frames, the feature vectors are stacked into a sequence and processed by a bidirectional LSTM, allowing each frame to be scored in the context of the full sweep [3, 5]. A linear projection followed by a sigmoid produces one quality score in [0, 1] per frame. **Masking:** Variable-length sequences are padded and masked during temporal processing, loss calculation, and evaluation.

#### 3.3 Gaussian Soft Labels

For each sweep, annotators mark one or more key frames, the frames on which an ONSD measurement is valid. We denote the indices of these key frames as *K* = {*k*_1_, …, *k*_*m*_} for a given sweep (*m* may be zero when a sweep contains no valid view). Rather than converting K into a hard 0/1 per-frame label, we construct a continuous soft label that encodes graded frame quality around each key frame.

#### Soft-label construction

Each key frame *k ∈ K* induces a Gaussian curve, and the per-frame soft label *y*_*t*_ is the maximum over all keys:

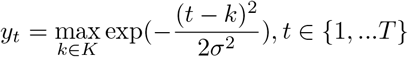

with *y*_*t*_ = 0 for every frame when K is empty (a no-key sweep). The width *σ* controls how quickly quality decays away from a key; we fix *σ* = 5 frames a priori in all experiments. Inter-annotator agreement was not evaluated, so *σ* was not calibrated to annotator variability; however, a post-hoc sensitivity analysis (Table 3) confirms that the performance was stable across the tested *σ* values. Taking the maximum rather than a sum keeps *y*_*t*_ in [0, 1] and prevents nearby key frames from inflating the target, yielding a value of 1 at each labeled key frame that decays smoothly with temporal distance.

### 3.4 Experimental Setup

#### Data splits

We used five-fold subject-level cross-validation over 18 subjects. The held-out folds contained 4, 4, 4, 3, and 3 subjects, with the corresponding training folds containing 14, 14, 14, 15, and 15 subjects. All sweeps from each subject were assigned exclusively to one fold, and results were aggregated over predictions from all held-out folds. No separate test set was used. **Baselines:** Following prior ultrasound frame-comparison work [7], we compare against Root Mean Squared Error (RMSE) and Normalized Cross Correlation (NCC) as pixel-wise baselines. Within each fold, every held-out frame is compared with all key frames from the training subjects. We assign each frame its best-match value: the minimum RMSE or maximum NCC across the training key frames. We similarly use the maximum cosine similarity in frozen USFM feature space. Frames are ranked by increasing RMSE or decreasing NCC and cosine similarity. We also evaluate temporal hard-label models trained with either a frozen or fine-tuned encoder. **Ablations:** We evaluate sensitivity to *σ ∈* {3, 5, 7, 9} and remove the LSTM to test frame-independent scoring; the USFM encoder remains frozen in both experiments. **Evaluation metrics:** Evaluation is performed on sweeps containing at least one valid frame. Hit@1/3/5 measures whether a labeled key frame appears among the top-*k* predictions. We also report Mean Minimum Distance (MMD), the distance from the top-ranked frame to the nearest labeled key frame.

### 3.5 Implementation, Compute Resources, and Inference Time

We used a frozen USFM encoder (ViT-B, *D* = 768). The temporal head was trained for 200 epochs using Adam, a learning rate of 5 *×* 10^*™*5^, a batch size of 8, and masked binary cross-entropy loss. Experiments were performed on an NVIDIA Tesla V100 GPU. USFM frame embeddings were computed once and cached, after which only the bidirectional LSTM and linear scoring head were trained. End-to-end inference was measured with a sweep batch size of one and included DICOM loading and decoding, frame preprocessing, frame-wise USFM feature extraction, and temporal scoring. The current implementation achieves inference time of 3.2 *±* 0.3 seconds per sweep on an NVIDIA Tesla V100 GPU and 40.1 *±* 2.4 seconds per sweep on a compute node equipped with an Intel Xeon Gold 6130 CPU. Values are reported as mean *±* standard deviation.

## 4. Results

Table 1 compares the overall performance of the proposed method with the baselines. H@k is the percentage of sweeps containing a labeled key frame among the top-*k* predictions; MMD is the distance from the top-ranked frame to the nearest ground truth key frame. Our approach outperforms the baselines across all metrics. Table 2 reports a breakdown of performance for different acquisition views. H@1 is reported as a percentage and MMD in frames. Table 3 reports sensitivity to *σ*. The proposed method achieves 82.2% H@1 and an MMD of frames. This improves H@1 over hard-label with frozen encoder (71.9%) and the strongest training-free baseline (49.2%). Fine-tuning substantially underperforms frozen-feature learning. Per-view results are reported in Table 2. The soft-label model matched or exceeded the hard-label model in H@1 across all nine acquisition views, with the largest gains for leftward translation and upward fanning. MMD improved for most views but increased for upward translation and downward fanning.

**Table 1.** Five-fold evaluation of key frame selection. Our approach outperforms the baselines across all metrics. Best values for each metric are shown in bold.

| Approach | H@1 ↑ | H@3 ↑ | H@5 ↑ | MMD ↓ |
| --- | --- | --- | --- | --- |
| Random | 33.1% | 68.2% | 78.5% | 20.1 |
| RMSE | 33.9% | 38.8% | 45.5% | 13.6 |
| NCC | 30.1% | 35.1% | 39.2% | 18.3 |
| Cosine Similarity | 49.2% | 61.6% | 66.5% | 14.4 |
| Hard-label model (Frozen Encoder) | 71.9% | 75.6% | 78.9% | 3.89 |
| Hard-label model (Fine-tuning) | 52.9% | 63.2% | 68.6% | 9.46 |
| Soft-label model (Ours) | **82.2%** | **84.3%** | **86.0%** | **3.07** |

**Table 2.** Performance by acquisition view for hard- and soft-label models. H@1 is reported as a percentage and MMD in frames; best values are bolded. Soft label approach outperforms the hard-label across majority of the views.

| Acquisition view | H@1 ↑ | | MMD ↓ | |
| --- | --- | --- | --- | --- |
|  | Hard | Soft | Hard | Soft |
| Optimal Frame | **100.0** | **100.0** | **0.00** | **0.00** |
| Upward translation | **85.7** | **85.7** | **1.29** | 2.43 |
| Downward translation | 63.9 | **75.0** | 4.50 | **2.75** |
| Rightward translation | **100.0** | **100.0** | **0.00** | **0.00** |
| Leftward translation | 58.3 | **100.0** | 8.20 | **0.00** |
| Clockwise rotation | 55.6 | **69.4** | **4.86** | **4.86** |
| Counterclockwise rotation | **86.1** | **86.1** | **1.43** | **1.43** |
| Upward fanning | 41.7 | **61.1** | 14.80 | **1.80** |
| Downward fanning | **61.1** | **61.1** | **3.20** | 9.00 |

**Table 3.**
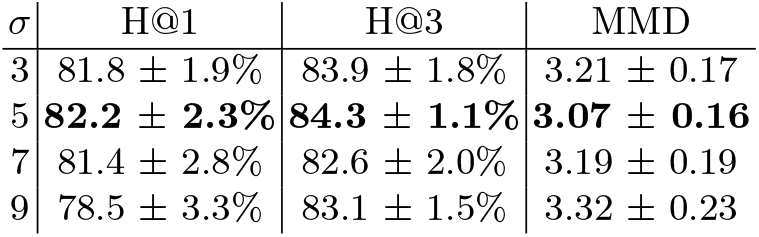
Performance across *σ* values over 5 folds. Values are reported as mean *±* standard deviation across the five folds. Similar performance across the tested *σ* values indicates that our approach is not too sensitive to the *σ* value.

| $\sigma$ | H@1 | H@3 | MMD |
| --- | --- | --- | --- |
| 3 | 81.8 ± 1.9% | 83.9 ± 1.8% | 3.21 ± 0.17 |
| 5 | **82.2 ± 2.3%** | **84.3 ± 1.1%** | **3.07 ± 0.16** |
| 7 | 81.4 ± 2.8% | 82.6 ± 2.0% | 3.19 ± 0.19 |
| 9 | 78.5 ± 3.3% | 83.1 ± 1.5% | 3.32 ± 0.23 |

Figure 2 shows predicted score trajectories for two representative sweeps. The left example contains a broad labeled valid-frame span, whereas the right example contains a narrower labeled span. In both cases, the soft-label model produces a smoother temporal response concentrated within the valid region than the hard-label model. Figure 3 provides a frame-level example from a single sweep. The soft-label model scores both the boundary and interior key frames highly, whereas the hard-label model assigns a substantially lower score to the boundary frame; both models suppress the later non-key frame.

**Fig. 1.**
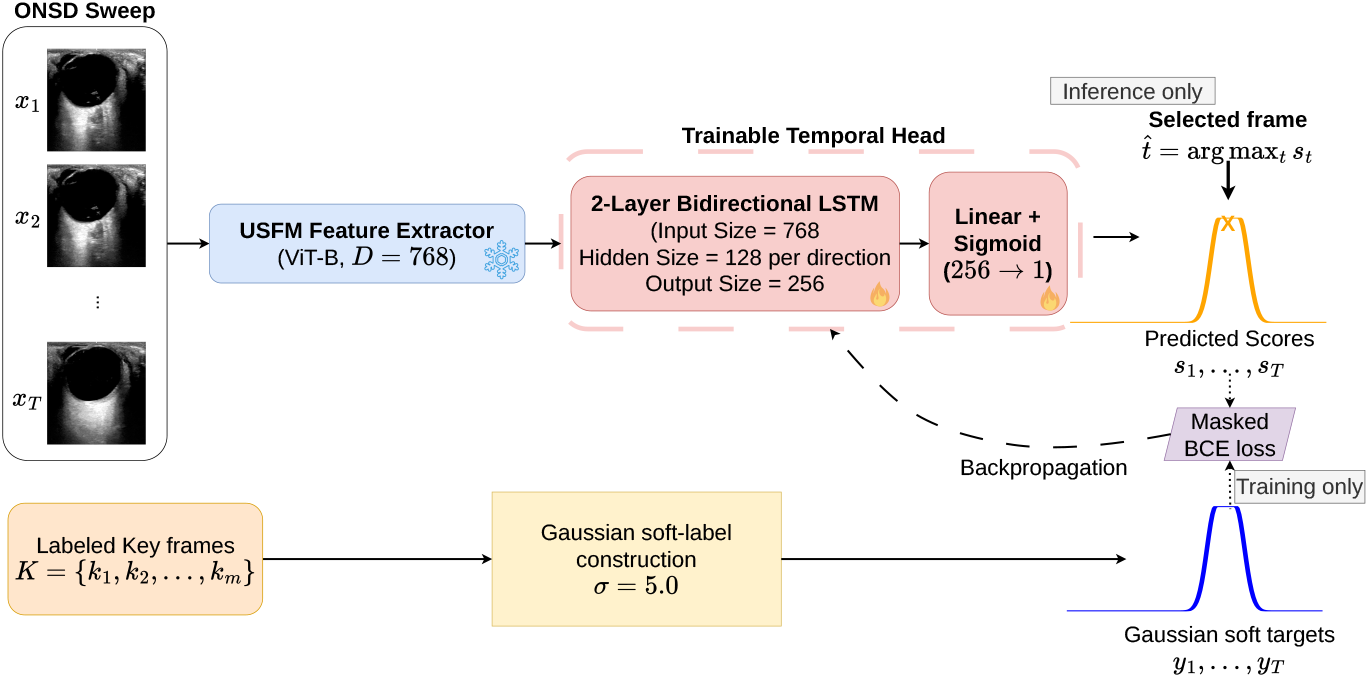
ONSD frame-quality scoring framework. A frozen USFM encoder and temporal head produce per-frame scores. Gaussian soft targets from labeled key frames supervise the temporal head during training, while inference selects the maximum-scoring frame.

**Fig. 2.**
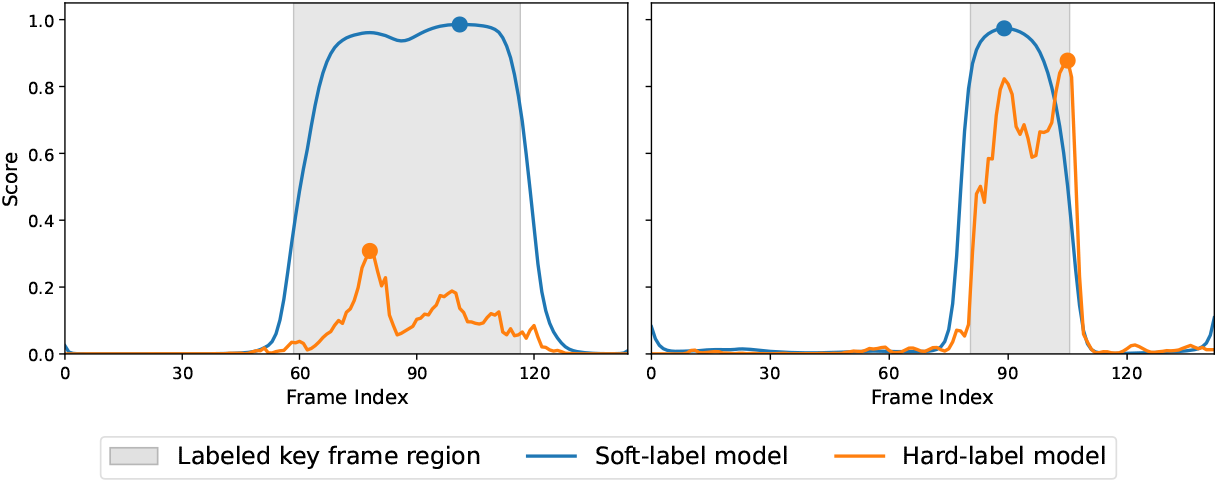
Predicted frame-quality scores for two representative ONSD sweeps. Shaded regions indicate labeled valid frames, and markers denote the top-ranked frame selected by each model. The soft-label model produces smoother temporal responses concentrated within the valid region than the hard-label model.

**Fig. 3.**
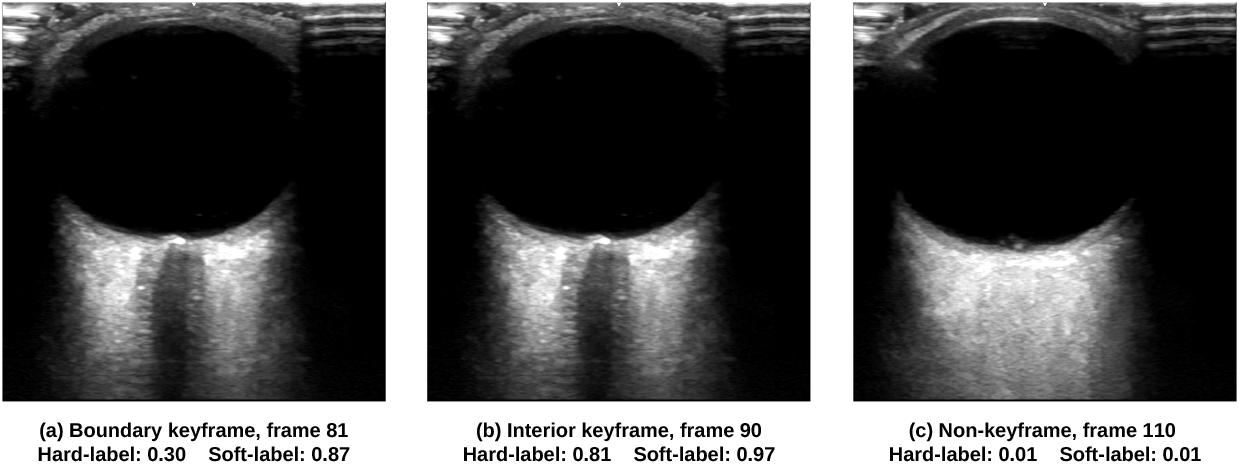
Frame-level predictions from one ONSD sweep. The soft-label model assigns high scores to boundary and interior key frames, while both models assign a low score to the non-key frame at frame 110, where the optic nerve sheath is not clearly visible.

### Ablation studies

We conducted several ablation studies. Removing the temporal head reduced H@1 to 74.4% and increased MMD to 3.76, indicating the sequence model contributes beyond per-frame features. We also evaluated four values for *σ* across five folds. Table 3 provides the results for this ablation. Performance remained consistent across the four *σ* values tested, indicating the method is not sensitive to the choice of label width.

## 5 Discussion

We presented a sparsely supervised framework for ONSD key frame selection that combines Gaussian soft labels with temporal scoring over frozen ultrasound foundation-model features. The method selected a key frame in 82.2% of evaluated sweeps, compared with 49.2% for the strongest training-free baseline and 71.9% for the hard-label model. The hard-label comparison and temporal ablation further indicate that both soft labels and temporal information contribute to the improved performance. Our work has limitations. The evaluation is based on a small, single-site cohort of 18 subjects, limiting conclusions about generalizability across subjects, scanners, operators, and sites. In addition, localization metrics exclude the 80 sweeps that do not contain any key frame, and the ability to reject such sweeps at inference was not evaluated. H@1 and MMD are more discriminating metrics in this study because learned methods tend to rank adjacent frames together. Random selection at H@3 and H@5 performs well because independent samples cover more of the sweep. We note that end-to-end fine-tuning underperformed frozen-feature training in this small cohort, although the cause of this difference was not investigated. Also, this work evaluates frame selection only and does not establish improvement in downstream ONSD measurement accuracy or clinical decision-making. Overall, the proposed sparsely supervised framework selects measurement-valid ONSD frames using only sparse labels and outperforms the hard-label model (82.2% vs. 71.9%), supporting graded targets that preserve relative frame quality near ambiguous valid-region boundaries rather than forcing abrupt binary decisions. Future work will evaluate multi-site generalization and integrate frame selection with auto-mated ONSD measurement.

## Data Availability

The de-identified ultrasound data used in this study are not publicly available due to patient privacy and institutional data-use restrictions. Data may be made available upon reasonable request, subject to institutional approval and applicable data-use agreements.

## 6 Impact in Resource-Constrained Settings

This work is the first step towards providing automated guidance for ultrasound acquisition, automated quality assessment, and objective ONSD measurements in low resource settings. The overarching goal is to enable medical personnel with limited ultrasound experience to perform reliable neurological examinations with greater consistency. The proposed framework reduces two important resource demands: the scarcity of labeled ONSD data and the scarcity of experts. We used an ultrasound foundation model to mitigate the limited ONSD data available while still achieving good results. Our framework only requires sparse frame-level labels rather than segmentation masks and our model supports CPU-only inference, although further optimization may be required for real-time use.

## Notes

### Competing Interest Statement

The authors have declared no competing interest.

### Author Declarations

The Institutional Review Board of Baylor College of Medicine gave ethical approval for this work (protocol ID: H55281)

